# Child and family experiences of a whole-system approach to physical activity: a citizen science evaluation protocol

**DOI:** 10.1101/2022.10.18.22281188

**Authors:** Marie Frazer, Amanda Seims, Michael J Tatterton, Bridget Lockyer, Daniel D Bingham, Sally E Barber, Andy Daly-Smith, Jennifer Hall

## Abstract

**Introduction:** Whole systems approaches are being adopted to tackle physical inactivity. The mechanisms contributing to changes resulting from whole system approaches are not fully understood. The voices of children and families that these approaches are designed for need to be heard to understand what is working, for whom, where, and in what context. This paper describes the protocol for the children and families’ citizen science evaluation of the JU:MP programme, a whole systems approach to increasing physical activity in children and young people aged 5-14 years in Bradford, UK.

**Methods and analysis:** The evaluation aims to understand the lived experiences of children and families’ relationship with physical activity and participation in the JU:MP programme. The study takes a collaborative and contributory citizen science approach, including focus groups, parent-child dyad interviews and participatory research. Feedback and data will guide changes within this study and the JU:MP programme. We also aim to examine participant experience of citizen science and the suitability of a citizen science approach to evaluate a whole systems approach. Data will be analysed using Framework approach alongside iterative analysis with and by citizen scientists in the collaborative citizen science study.

**Ethics and dissemination:** Ethical approval has been granted by The University of Bradford: Study One (E891-focus groups as part of the control trial, E982-parent-child dyad interviews), Study Two (E992). Results will be published in peer-reviewed journals and summaries will be provided to the participants, through schools or directly. The citizen scientists input to create further dissemination opportunities.

**Article Summary:** *Strengths and Limitations of this study:*

- This protocol is the first, to our knowledge, to describe a citizen science-based evaluation of a whole systems approach to physical activity with children and families.
- The novel and innovative study design allows children and families to be at the centre of our understanding of what encourages and discourages them to be active.
- By conducting citizen science as part of a reactive process evaluation, improvements to the research and the implementation can be made in real time, centred around those who matter most
- The study emphasises the importance of the research participant experience within citizen science and sets out how to evaluate and improve experience.
- Limitations include a small sample size. Whilst this is intentional as it will allow us to capture in depth, meaningful data over time, it will likely make it more difficult to capture a diverse range of experiences. There is an option for the children participating in the collaborative citizen science study to conduct research amongst their wider peers, this is dependent on whether they want to do this, to allow them freedom and ownership over the research

## 1.0 Introduction

### 1.1 Physical activity levels and impacts

Across Europe, less than a third of 2-18 year olds achieve the recommended 60 minutes of moderate-to-vigorous physical activity per day (1). Such high levels of physical inactivity are fuelling a worldwide public health problem and negatively impacting children’s physical fitness, cardiometabolic health, bone health, cognitive outcomes, mental health and adiposity (2, 3). Physical activity levels reduced further during the Covid 19 pandemic (4), with recent evidence suggesting the reductions have remained post-lockdown (5). Children from socially disadvantaged areas and ethnic minorities - who were already the least active - were most negatively impacted by the covid pandemic, further exacerbating health inequality (6-9). Additionally, across all age groups, girls are less active than boys, creating gendered health inequity (1).

### 1.2 Progressing to Whole Systems Approaches to Physical Activity

To date, most interventions have focused on individual behaviour change, resulting in minimal effects on physical activity behaviours (10, 11). To address this issue, the World Health Organisation (WHO) Global Action on Physical Activity (GAPPA) report proposes a systems-based approach, involving cross-government, multi-sectoral partnerships and community engagement (12). To enact effective systems-based approaches, it is recommended that programmes work closely with local people to develop solutions tailored to intended recipients’ context and experience (13-15). Recently, Sport England (the arms-length body of government responsible for growing and developing grassroots sport and getting more people active across England) has invested £100 million across 12 Local delivery pilots (12). The purpose of each pilot is to design and implement a whole-system physical activity approach, providing a unique opportunity for evaluation.

### 1.3 Evaluations of whole-system physical activity interventions

There is a growing body of evidence surrounding whole-systems evaluations (14, 16, 17) that suggests mechanisms underpinning complex and whole-systems interventions are likely more varied and dynamic than singular or less complex interventions (18). As physical activity interventions become more complex, it is increasingly important to explore what works, where, for whom and in what contexts (19). This is essential to understand the transferability, replicability and upscaling of interventions and to inform policy change (20). Additionally, timely and integrated evaluation can inform dynamic systems change through continuous improvement of the intervention design and implementation (14).

Literature reviews of evaluations of system approaches to health recommend several approaches and methods; the embedded researcher approach, qualitative inquiry (process evaluation) through a systems thinking lens, systems mapping, network mapping, ripple effects mapping, and dynamic systems modelling (14, 17, 21-23). Such methods are applied in evaluations of whole systems approaches ((14, 17, 19, 24)). However, to our knowledge, there are no existing guidelines, protocols or evaluation studies reporting on evaluating whole-systems approaches focussed on children and families. Given the increasing adoption of whole systems approaches to physical activity and/or other health behaviours, there is a need to build further evidence around appropriate, effective, and innovative methods for evaluating systems-based interventions (21, 22), including those involving children and families.

### 1.4 Using citizen science to evaluate whole system approaches to physical activity

The United Nations asserts that children have the right to contribute to decisions that impact them personally and affect the services they use(25). Therefore, it is essential that children and their families are integral stakeholders within the evaluation of whole-system physical activity approaches. Moreover, including local people promotes a more comprehensive and contextual understanding of what works and for whom (14). At present, previous evaluations of whole system physical activity approaches have not placed children and families at the centre. One way to address this is through involving the public through a citizen science research approach, which could improve research quality and lead to system changes (26). By placing children and families at the heart of research (27, 28), their needs can be better understood, and programmes adapted accordingly. A growing cross-disciplinary body of evidence demonstrates the benefits of taking different citizen science approaches with young people (29-35). Citizen science has been used successfully to understand young people’s physical activity experiences (36). However, the potential to understand whole systems physical activity approaches using citizen science has not been realised (22).

A key principle of citizen science is that citizen scientists should benefit from participating (37). Research indicates that positive experiences for young people can be achieved by considering power dynamics, relationships, and personal growth within citizen science (29, 32, 38-40). Furthermore, it is recommended that citizen science projects evaluate participant experience to understand the value of young people’s contribution and to improve outcomes (41, 42). This purpose of this paper is to describe a citizen science evaluation approach of the Join Us: Move, Play (JU:MP) whole-system physical activity intervention with children and families. A secondary purpose is to outline the evaluation of participant experience within the citizen science process.

## 2.0 Methods and analysis

### 2.1 Aims and objectives

This paper describes a protocol for a citizen science research study, aiming to understand the mechanisms through which a whole-system approach to physical activity (JU:MP) influences behaviour change amongst families and to evaluate participant experience within this citizen science project. The specific objectives are:

1. To understand perspectives and lived experiences around the physical activity of children and families in JU:MP delivery areas
2. To assess the feasibility, fidelity, and acceptability of JU:MP among children and parents/carers
3. To examine the mechanisms of change that underpin the physical activity behaviour of children and families, when, how and why this happens within JU:MP
4. To contribute to dynamic systems change through informing programme refinement based on ongoing findings from objectives 1-3
5. To formatively understand children and families experience as participants within a contributory and collaborative citizen science approach, to inform continuous study delivery improvements

### 2.2 Study context

During the pandemic, 73% of Bradford children (9-13 years) were not meeting physical activity guidelines. On average, children of South Asian heritage and females were less active than their white British peers and males respectively (4). Within the Bradford district, the number of children overweight or obese is higher than the national average (37.9% vs 34.2%), with higher levels in the most deprived areas (43). Specifically, within the JU:MP area, average income is significantly below the UK average (44). Bradford is the youngest city within the UK, with 24% of residents under the age of 16 (43).

#### 2.2.1. Join Us: Move, Play (JU:MP)

JU:MP is a whole-system physical activity programme, being delivered in the north area of the city of Bradford, UK. Evidence-led co-production and formative evaluation inform ongoing intervention refinement. JU:MP consists of 15 work streams informed by ISPAH’s eight investments that integrate physical activity across various sectors e.g., active travel, schools, and community ((45)). These work streams are aimed at creating societal (e.g., local authority policy), personal (e.g. how families travel to school), organisational (e.g. changes to the culture and environment of schools and Islamic Religious Settings (46, 47), and environmental (e.g. green space and parks) change. Neighbourhood action groups (across eight areas) consisting of local cross-sector stakeholders develop, tailor, and enact the JU:MP programme at a hyper-local level. Children and families could interact with or be influenced by any of the 15 work streams. JU:MP is being delivered in two phases across different neighbourhoods. Within three pioneer neighbourhoods (pathfinder phase: 2019-2023), the intervention is now embedded. Five accelerator phase neighbourhoods began the JUMP programme in 2021 with programme delivery continuing until 2024 (Figure 1). Further information can be found in Hall (2021)(19)

**Figure 1.**
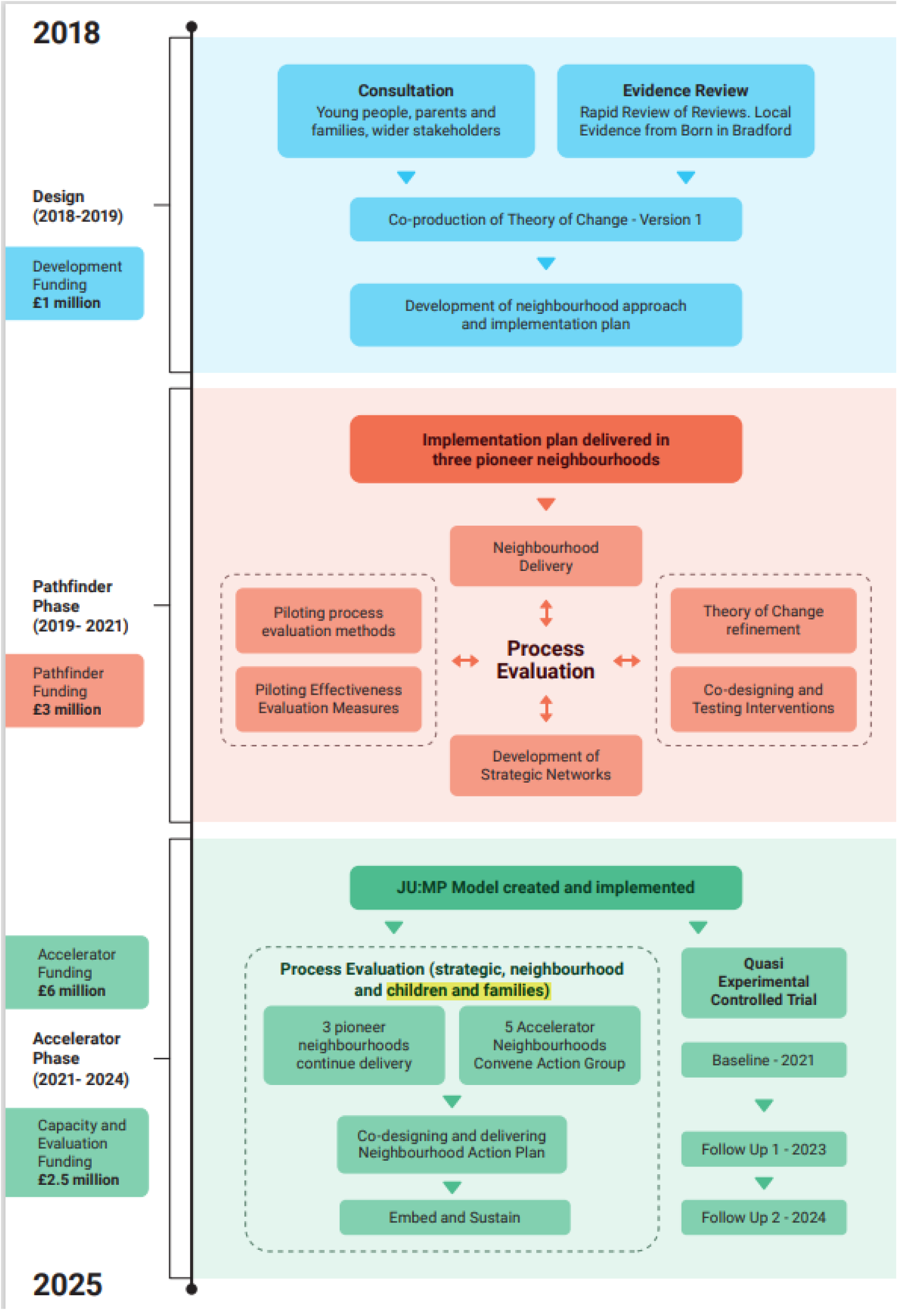
Timeline of the JU:MP program

#### 2.2.2 The overarching JU:MP evaluation

The JU:MP evaluation employs a mixed methods approach which sits within the complementary philosophies of realist and ‘systems thinking’ methodology (48). The concurrent mixed method approach contains two main elements: an effectiveness and a process evaluation (Figure 1). The effectiveness evaluation includes: (1) a controlled trial focused on primary-age children, examining effectiveness at the neighbourhood level, and (2) a pre-post evaluation of children within the Born in Bradford cohort study at age 7-11 (49), and again at age 13-15, examining effectiveness at the population (North Bradford) level. The primary outcome is children’s moderate-vigorous intensity physical activity (50).

The process evaluation includes an examination of the mechanisms and contextual factors influencing the implementation and impact of JU:MP and includes a focus on policy and strategy, overarching work streams, the JU:MP neighbourhood approach, and children and families. A mixed-method data collection approach includes semi-structured interviews, observations, documentary analysis, surveys and participatory evaluation methods (e.g. reflections and ripple effect mapping). For further information on the overarching process evaluation, see Hall et al. (2021) (19). This paper describes two citizen science evaluation studies that are part of the overarching process evaluation: 1. an interview and focus group study with primary-age children and their families and 2. a year-long collaborative study with secondary-age children and their families.

### 2.3 Patient and public involvement

Public involvement through the citizen science methodology is integral to this study, as described in the methods section of this protocol. Wider public involvement with youth research ambassadors from Born in Bradford (50) shaped the overarching study design, including for example incentives, methods, and realistic time commitments.

### 2.4 Citizen science approach

Two interlinked longitudinal citizen science studies will help understand child and family experiences of the JU:MP programme. These will be complemented by an evaluation of the citizen scientists’ experiences.

Citizen science is a trans-disciplinary participatory method (51). Whilst there is no agreed definition (52-54), for this paper, we define citizen science as the involvement of members of the public who work with professional scientists to advance research (55). Citizen science projects can be viewed on a continuum. On one end, citizens are ‘passive contributors’ to activities (56); on the other, citizen scientists are fully immersed in a local community, and the research is a joint enterprise to help identify and solve societal issues (56). Using Shirk et al.’s (57) typology, study one adopts a contributory model approach, where projects are designed by researchers and members of the public primarily contribute data. Study two adopts a collaborative model approach, where researcher staff create the project and ‘members of the public contribute data and help to refine project design, analyse data, and/or disseminate findings’ (57).

Guidance exists on conducting high-quality citizen science research (37), including young people in research ((58)) and co-production in a Bradford context (59). The following seven principles, adapted from the above documents, will guide the current citizen science approach:

1. Child-friendly involvement, including clear communication and feedback.
2. Power should be shared with the children where possible.
3. Voluntary participation should be offered at different stages of the research.
4. Researchers should respect the participants’ knowledge; one way of showing this respect is by going to the communities.
5. Research should be inclusive and relevant to the children’s lives.
6. Participation should include a reciprocal relationship where training supports participants.
7. Researchers should ensure that participants are safe and sensitive to risk. Research leads should consider ethical and legal issues.

A longitudinal research design will provide an understanding of families’ direct experience with JU:MP and how this evolves (60, 61). The longitudinal nature will facilitate the development of a meaningful relationship between the child, families, and the researchers (28, 29), provide time to explore change mechanisms and take seasonal variations in children’s physical activity levels and sedentary time into account (62).

### 2.5 Study design

Herein we describe the design of the two studies that make up the citizen science evaluation, including contributory citizen science (study one) and collaborative citizen science (study two); see Figure 2.

**Figure 2:**
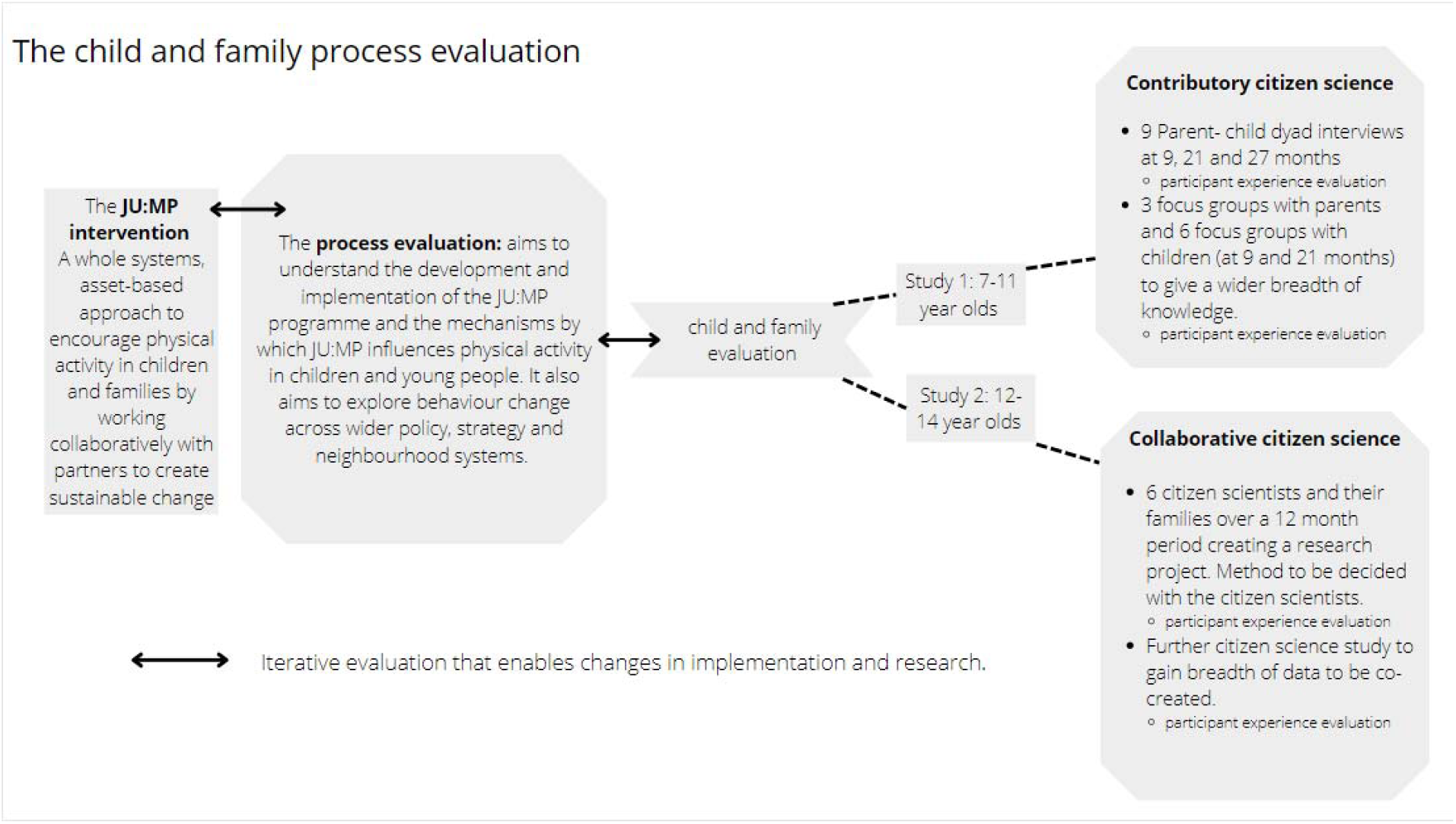
The child and family process evaluation

The two studies have been designed in a way that the level of citizen science participation required is age-appropriate (63-65). For the collaborative citizen science study (ages 12-14), participants choose the study method, which may involve independent data collection in different neighbourhood locations and/or independent use of technology. These methods may not be suitable for primary age children due to parental concerns of personal access to smartphones(66, 67). The contributory citizen science study will accommodate the views of younger children and their families through focus groups and interviews (64). Ongoing findings from both studies will inform JU:MP programme development and delivery through six-weekly research and implementation team meetings.

Ethics approval was granted by the Chair of Humanities, Social and Health Sciences Research Ethics Panel at the University of Bradford for both studies (June 2022). For study two, given the collaborative citizen science approach, ethics approval covers participant on boarding, the study in principle and the first workshop. Ethical amendments will be submitted at (at least) two further points: 1. once the data collection method has been decided on and 2. once the data analysis approach has been finalised (See table 2)

### 2.6 Study one - Contributory citizen science with primary-age children and their families

Study one uses two data collection methods; focus groups (separate child and parent focus groups) and parent-child dyad interviews. Focus groups allow for a diverse range of responses and provide children with less intimidating environments than interviews (64). The parent-child interviews - which pair a child with their primary caregiver - allow for more detailed and longitudinal exploration of people’s experiences and engagement with JU:MP, and physical activity behaviour change mechanisms. Parent-child dyad interviews will aid understanding of how parents shape children’s behaviour as well as allowing parents to expand on the child’s verbal expression (68, 69). The study draws on a range of concepts relevant to intervention evaluation that the focus group and interview topic guides were informed by (48); see table 1.

**Table 1.** Concepts and their application within the study.

| Concept | Definition | Application within the study |
| --- | --- | --- |
| Feasibility (70) | Feasibility: How easy is it to participate in different elements of JU:MP | ☐ what has made it easy/hard to engage with JU:MP<br>☐ what would make it easier to engage with JU:MP |
| Participant fidelity (71-73) | The extent to which participants understand the concepts and purpose of the intervention and are exposed to and engage with the intervention. | ☐ exposure participants have to different elements of JU:MP<br>☐ receipt of knowledge of physical activity and its benefits; and knowledge and understanding of the JU:MP intervention.<br>☐ responsiveness - the extent to which participants feel the various components of JU:MP are useful<br>☐ engagement - the components they report seeing/hearing about and/or engaging with/ or taking part in |
| Acceptability (74) | Anticipated or experienced | ☐ Affective attitude (anticipated or experienced thoughts and feelings about JU:MP) |
|  | cognitive and emotional responses to the intervention. | ❑ burden (the anticipated or experienced amount of effort required to engage with JU:MP)<br>❑ ethicality (the extent to which JU:MP fits within their value system)<br>❑ self-efficacy (participant's confidence that they can perform the behaviour required to participate in JU:MP)<br>❑ opportunity costs (anticipated or experienced benefits or values that are given up to engage with JU:MP) |
| Mechanisms of change (75) | How does the intervention produce change within the delivery context | ❑ what elements of JU:MP works, for whom, and in what context<br>❑ wider intended or unintended impacts of JU:MP |

#### 2.6.1 Study one: Sampling, recruitment, and incentives

Focus group and interview participants will be recruited from the three neighbourhoods included in the JU:MP control trial (see section 2.2.2) to permit integration of findings across evaluation components. Each family will receive a £20 voucher per interview as a token of appreciation for their time.

##### Focus group recruitment

parents and children will be recruited from the sample who provided consent and assent to engage in the JU:MP control trial when in primary years one to three (aged five to eight). The sample for the focus groups will be recruited when the children are in years two to four (aged six to nine). Schools will be selected from different areas to ensure a geographical spread of participants. Parent focus groups will typically take place in different schools to child focus groups to enhance the diversity of responses. Children and parent participants will initially be randomly selected but revised based on advice from teaching staff (e.g., if the family has since left school, or if the sample lacks diversity in socioeconomic backgrounds, ethnicity, or physical activity behaviours). Different participants will be selected at different data collection time points to allow a range of families to share their experiences. Two weeks before the child focus group, parents will receive a letter or email from their child’s school to notify them and provide an opportunity to withdraw consent. Verbal consent (parent group) and assent (child focus group) will be recorded at the start of each focus group.

##### Parent-dyad interviews

the eligibility criteria are: 1. live in the JU:MP control trial neighbourhoods and 2. include a child in primary years two, three or four. Participants will be recruited through local community networks and social media. A random sample will be selected from those who express interest in participating and have suitable availability. Informed verbal consent will be obtained from adult participants, and assent from child participants.

#### 2.6.2 Study one: Data Collection Methods

Interview data collection will take place approximately nine, 21 and 27 months after the JU:MP accelerator-phase programme launch (Sept/Oct 2022, Sept/Oct 2023 and Mar/Apr 2024). Focus group data will be collected at the first two time points. Multiple data collection time-points enables findings to provide repeated feedback on programme delivery to enhance the likelihood of engagement with and impact of JU:MP. Participants’ basic demographics (postcode district/name of neighbourhood; gender; age; ethnic group) will be collected during the interviews and focus groups to describe the sample characteristics relative to the population.

##### Focus groups

Six focus groups with children and three focus groups with parents will be held in schools. Two researchers will be present at each focus group; one to facilitate the session and one to take reflexive field notes. The children’s focus groups will incorporate the Write, Draw, Show & Tell (76) technique to explore children’s understanding of physical activity. This participatory visual method allows children to express their views, thoughts and emotions non-verbally and/or verbally, facilitating inclusivity and engagement (76). Images depicting various components of JU:MP will also facilitate discussions. See table 1 for an overview of focus group discussion topics.

Parent-child dyad interviews: Nine parent-child dyads (three from each neighbourhood) will be interviewed at each time point. Photo elicitation methods will be used to stimulate discussion. Before each interview, families will be asked to take photos showing their physical activity as part of a normal week and images depicting various components of JU:MP will be shown in interviews. Both sets of images will be used to facilitate discussions around child and family physical activity behaviours. Interview questions will focus on the parent and child’s understanding of physical activity, their (change in) physical activity behaviours, why they may or may not have engaged with the JU:MP programme and what impact this has had (table 1).

#### 2.6.5 Study one: Data analysis

Data analysis will be undertaken using a framework approach (77). Interviews and focus groups will be audio-recorded and transcribed verbatim. Once imported into NVivo (QSR International, Melbourne, Australia), data will be coded into a framework previously developed to analyse wider JU:MP process evaluation data. This will facilitate the integration of data and findings with the broader process evaluation. Further detail as to how the framework was developed and the rationale for this approach can be found in Hall *et al*. (19). A recurrent cross-sectional analysis will be applied for the focus group data to explore differences across the three different focus group cohorts(51). For interview data, a trajectory analysis will be used to examine change over time within the parent-child dyads participating in the study.

### 2.7 Study two: Collaborative citizen science with secondary school-aged children and families

Study two is a collaborative approach - with data collection developed in partnership with the citizen scientists. The longitudinal study will take place over a year (Sept 2022-Sept 2023) and focus on the citizen scientists and their families experiences of physical activity and the JU:MP intervention.

#### 2.7.1 Study two sampling recruitment and incentives

We aim to recruit six female citizen scientists, aged 12-14, and their families. We are targeting females due to females being consistently less active than males (1) and our public involvement work indicating that families, particularly those from South Asian backgrounds, may be more accepting of a single-sex research project. Participants will be selected to ensure a diverse sample according to family composition, ethnicity, geographical location, and previous interaction with JU:MP. Three families will be selected from one pioneer neighbourhood (60% Pakistani ethnicity) and three from an accelerator neighbourhood (75% White British ethnicity), enabling exploration of families’ experiences at different stages of the JU:MP delivery process. The household earnings in both areas are below the national average (44), and the researchers have taken steps to ensure socioeconomic status should not be a barrier to recruitment.

Families will be approached through JU:MP Community Engagement Managers (CEMs) and other partners who work directly with the communities, to facilitate trust (30, 78). Informed consent from the parents and assent/consent from the children will be obtained before data collection commences and at key data collection points as the project evolves.

The study two incentives package was developed in partnership with the youth research ambassadors from Born in Bradford prior to the commencement of the study. The incentives (to the total value of £400 per family) given throughout the study include family vouchers, child vouchers, money donated to the child’s charity of choice and team-building activities.

#### 2.7.2 Study two: Data collection methods

Over the longitudinal study, the citizen scientists will be expected to collect data every fortnight on personal and family experience of JU:MP and physical activity. The citizen scientists will attend training workshops on how to conduct ethical research, plan and carry out data collection and analysis. Families will be asked to contribute to the data collection and, if the citizen scientists see it as appropriate, comment on the data analysis.

There will be six group workshops in addition to regular contact between the researcher and citizen scientists across the year (See table 2). The first two workshops will be facilitated by a researcher, and subsequent workshops may be co-led with citizen scientists as the project develops. The first workshop will be held in a location mid-way between the recruitment localities. Citizen scientists will be asked if this is a suitable location for future workshops.

**Table 2.** workshop plan

| Month | Schedule |
| --- | --- |
| 2022-2023 |  |
| Sep | **Workshop 1 – Full day**<br>Researcher led<br>• Research question agreed<br>• Data collection method selected<br>• Data analysis plans discussed<br>• Ethical code produced<br>• Timings of further meetings and data collection and the meeting location will be arranged.<br>*Participant evaluation*<br>**Ethical amendments** submitted- Once approved new information will be sent to families to consent |
| Oct | Data collection begins (Data collection every 2 weeks. Approx time spent by families is 20 min every two weeks -to be confirmed with CS.).<br>Meeting/contact with the CS to decide on final data analysis plans<br>**Ethical amendments** will be submitted now the data analysis method has been selected. |
| Nov | **Workshop 2 – ½ day**<br>• Researcher led<br>• Reflection on the process<br>• Data analysis training<br>• Data analysis with citizen scientists<br>• Families updated by citizen scientists<br>*Participant evaluation. Activity arranged - Citizen Scientists' choice. Family voucher* |
| Dec |  |
| Jan | Longer researcher check in to ensure Citizen scientists are supported |
| Feb | **Workshop 3 – ½ day**<br>• This and further workshops could be co-led with the citizen scientists.<br>• Data analysis<br>• Discuss further research with peers<br>• Discuss data analysis with families<br>• Family update by the citizen scientists<br>*Participant evaluation. Family and child voucher* |
| Mar | **Optional workshop to discuss peer citizen science study** |
| Apr |  |
| May | **Workshop 4**<br>• Team building activity<br>• Data analysis<br>• Decide next steps for the research - data dissemination including non traditional formats.<br>*Participant evaluation Activity arranged - Citizen Scientists choice. Family voucher* |
| Jun |  |
| 2022-2023 |  |
| Jul | **Workshop 5**<br>• Team building activity<br>• Data analysis<br>• Decide next steps for the research - data dissemination including non-traditional formats.<br>*Participant evaluation. Family and child voucher* |

Workshops one and two will focus on training and project development. Key decisions will focus on: finalising the research question, selecting the data collection method(s), ethical issues, how the citizen scientists want to transfer data, data analysis and dissemination. The citizen scientist will have the opportunity to use different methods, surveys, interviewing, photovoice and journaling within the workshop to inform their chosen data collection method. In later workshops, if appropriate, families will be brought into the workshops to see the data analysis. The workshops have been designed to be both accessible and engaging for children (79).

#### 2.6.3 Optional study

Citizen scientists will have the option to design and conduct a wider research study to understand physical activity experiences amongst their peers (of all/any gender). There is potential for the citizen scientists to gain breadth of data, and unique insights, given evidence that young people can be more open and willing to share experiences with peer researchers than professionals (80). This optional study will be introduced in workshop three, and if the citizen scientists want to take it forward, they will guide the study’s scope, reach and audience. Through this, the citizen scientists will learn further skills on how to plan and deliver a research project from the start, including first-hand experience of the academic ethics process.

#### 2.6.4 Study two: Data analysis

Citizen scientists will be introduced to different analytic approaches appropriate to the data collection method(s) selected, and will be supported to decide on an analysis method based on the options presented and their own ideas, and to undertake analysis of the data. Given the onerous nature of traditional data analysis approaches, such as thematic analysis (81, 82) citizen scientist and their families’ analysis is likely to take an adapted approach, such as producing stories or case studies from the data. We anticipate that citizen scientists will be involved in analysing data collected in the period in between workshops, from workshop 3 onwards, taking a cross-sectional approach.

Alongside the collaboratively chosen cross-sectional data analysis, data will also be analysed using a trajectory approach; this approach has been decided in advance by the researchers to take full advantage of the longitudinal data. It is adapted from the idea of rivers of multilingual reading (83).

The citizen scientists will choose which data is to be included and how. At each workshop, selected data will be placed along the ‘river’ and the families will note links or changes over time. This process of displaying the snapshots from each wave together will allow for identifying critical moments (84).

### 2.7 Overarching participant experience evaluation

The research team will evaluate the process and impact of the citizen science approaches on participant experience across both studies (85). The guidelines adopted in this research intended to promote a positive participant experience, outlined in section 2.3 (37, 58, 59), inform the evaluation. Three data collection methods will be used; a body sort exercise, an individual reflection card and researcher observations. Any adaptations to the citizen science process suggested by participants and/or researchers, and agreed upon by the participants, will be implemented on an ongoing basis.

The body sort exercise(79, 86) will take the form of a workshop activity with the citizen scientists. Cards will prompt key participant experience elements (e.g. power relations, inclusivity) in an age-appropriate format. Participants will be able to add further cards detailing other aspects of their experience. Participants will place cards representing their experience on the body outline, and engage in discussion around this. Within study one (section 2.5), the body sort exercise will occur at the end of three of the six children’s focus groups, and parent and child dyad participants will be asked to attend a focus group to discuss their interview experience, at each time-point. Within study two (see section 2.6), participant experience will be evaluated at the end of each workshop and will encompass citizen scientists’ experience since the last workshop. Citizen scientists will also have the option to complete an individual anonymised reflection card in case they do not want to voice their opinion in front of the group. A secondary researcher will collect participant experience data to mitigate social desirability bias (87).

Researcher observations, which allow an independent record of events and behaviours to be captured in real time (19), will be conducted to inform evaluation of participant experience. Researchers will observe focus groups (study 1) and workshops (study 2). Observations will not occur within the parent-child dyad interviews (study 1) to avoid creating a power imbalance between the researchers and the participants. The lead researcher will complete a reflexive research journal, focused on participant experience and improving our research to create a better participant experience (88).

## Data Availability

The dataset that will be generated and analysed during the current study are not publicly available to reserve the anonymity of research participants.

## 3.0 Ethics and dissemination

The current study adopts and advocates for a bespoke ethical approach for collaborative citizen science projects. Training for the participants, alongside a flexible, responsive approach to changes made as the project develops ensures the projects meet high ethical standards (88). Ownership of the research is an important ethical concept (30). Where information is to be disseminated, work will be credited to the citizen scientists while ensuring the participants’ and their family’s views on anonymity are respected. The collaborative citizen scientists are of an age where evidence suggests they can carry out a research project and express their expert knowledge (29, 88-90). However, as researchers under the age of 16, their consent alone is not legally adequate, and parent or guardian consent must be provided prior to research participation. Given the status granted to the citizen scientists, it seems incongruous not to allow them to provide their consent to take part in the project (86). As a result, to meet institutional ethical standards, both the children (citizen scientists) and their families will be required to give informed consent before any child’s participation in the research.

A key element of citizen science is acknowledging the voluntary contributions to research by citizen scientists, with debates on how this should be achieved (37, 91). Within both studies, incentives are used to compensate participants for their time. In line with ESCA’s characteristics of citizen science, incentives differ according to the project context and type (92). One of the key characteristics of citizen science is that it is a voluntary undertaking, and therefore there is a fine line between incentives and payment, which has been carefully considered (92). There is an emerging call for diversity in citizen science projects, with socioeconomic status being a known barrier to participation (93). To reduce the participants’ economic situation as a participation barrier, travel will be kept to a minimum, with research being conducted locally and any necessary travel costs reimbursed. Within study two, if technology is a barrier to a family participating, JU:MP will provide reasonable equipment (e.g., a tablet). The progress and findings of the study will be communicated to the citizen scientists and families in various ways, e.g., animated videos, and citizen scientists in study two will contribute to wider dissemination of study findings. Results will be published in peer-reviewed journals and summaries will be provided to the participants, through schools or directly.

## 4.0 Abbreviations

JU:MP: Join Us: Move. Play
LDP: Local Delivery Pilot
CEM: Community Engagement Manager

## Author contributions

This study is part of the evaluation package of the JU:MP whole-systems physical activity project in Bradford. SB and ADS led the development of the overarching JU:MP research design in partnership with JH, DB and AS. The conception and design of the studies presented in this paper was led by AS (study one) and MTF (study two). * JH and ADS provided equal senior author support for both studies, ensuring the alignment with the overarching process evaluation, with feedback from all authors. MTF led the writing of the initial manuscript with significant input from AS, JH and ADS. Subsequent drafts were commented on by all authors, and revisions were made by MTF. All authors have approved the submission.

## Ethics approval and consent to participate

Study One:- The University of Bradford (E891-focus groups as part of the control trial, E982-parent-child dyad interviews)

Study Two: - The University of Bradford (E992)

## Acknowledgements

We are grateful for the funding provided by Sport England to implement and evaluate the programme. The authors wish to thank the JU:MP implementation team and wider stakeholders, including expert colleagues within Born in Bradford and ActEarly, for their input in developing the child and family process evaluation and their ongoing engagement with and openness to the evaluation process. Thank you for the time and ideas of the young research ambassadors from Born in Bradford.

## References

1. Steene-Johannessen J, Hansen BH, Dalene KE, Kolle E, Northstone K, Møller NC, et al. Variations in accelerometry measured physical activity and sedentary time across Europe – harmonized analyses of 47,497 children and adolescents. International Journal of Behavioral Nutrition and Physical Activity. 2020;17(1).

2. Bailey R, Hillman C, Arent S, Petitpas A. Physical activity: an underestimated investment in human capital? J Phys Act Health. 2013;10(3):289–308.

3. Bull FC, Al-Ansari SS, Biddle S, Borodulin K, Buman MP, Cardon G, et al. World Health Organization 2020 guidelines on physical activity and sedentary behaviour. British Journal of Sports Medicine. 2020;54(24):1451–62.

4. Bingham DD, Daly-Smith A, Hall J, Seims A, Dogra SA, Fairclough SJ, et al. Covid-19 lockdown: Ethnic differences in children’s self-reported physical activity and the importance of leaving the home environment; a longitudinal and cross-sectional study from the Born in Bradford birth cohort study. International Journal of Behavioral Nutrition and Physical Activity. 2021;18(1):117.

5. Salway R, Foster C, de Vocht F, Tibbitts B, Emm-Collison L, House D, et al. Accelerometer-measured physical activity and sedentary time among children and their parents in the UK before and after COVID-19 lockdowns: a natural experiment. Int J Behav Nutr Phys Act. 2022;19(1):51.

6. Craike M, Wiesner G, Hilland TA, Bengoechea EG. Interventions to improve physical activity among socioeconomically disadvantaged groups: an umbrella review. International Journal of Behavioral Nutrition and Physical Activity. 2018;15(1).

7. Drenowatz C, Eisenmann JC, Pfeiffer KA, Welk G, Heelan K, Gentile D, et al. Influence of socio-economic status on habitual physical activity and sedentary behavior in 8-to 11-year old children. BMC Public Health. 2010;10(1):214.

8. Pampel FC, Krueger PM, Denney JT. Socioeconomic Disparities in Health Behaviors. Annual Review of Sociology. 2010;36(1):349–70.

9. Poulain T, Vogel M, Sobek C, Hilbert A, Körner A, Kiess W. Associations Between Socio-Economic Status and Child Health: Findings of a Large German Cohort Study. Int J Environ Res Public Health. 2019;16(5):677.

10. Mackay K, Quigley M. Exacerbating Inequalities? Health Policy and the Behavioural Sciences. Health Care Analysis. 2018;26(4):380–97.

11. Messing S, Rütten A, Abu-Omar K, Ulrike U-R, Goodwin L, Burlacu I, et al. How Can Physical Activity Be Promoted Among Children and Adolescents? A Systematic Review of Reviews Across Settings. Frontiers in Public Health. 2019;7:1–15.

12. World Health Organization (WHO). Global action plan on physical activity 2018–2030: more active people for a healthier world. Geneva; 2018.

13. Allender S, Occhipinti J-A, Bauman A, Bellew B, Cavill N, Chau J, et al. Getting Australia Active III: A systems approach to physical activity for policy makers 2020.

14. Potts A, Nobles J, Shearn K, Danks K, Frith G. Embedded Researchers as Part of a Whole Systems Approach to Physical Activity: Reflections and Recommendations. Systems. 2022;10.

15. Tibbitts B, Willis K, Reid T, Sebire SJ, Campbell R, Kipping RR, et al. Considerations for Individual-Level Versus Whole-School Physical Activity Interventions: Stakeholder Perspectives. Int J Environ Res Public Health. 2021;18(14):7628.

16. Sawyer A, Den Hertog K, Verhoeff AP, Busch V, Stronks K. Developing the logic framework underpinning a whole-systems approach to childhood overweight and obesity prevention: Amsterdam Healthy Weight Approach. Obesity Science & Practice. 2021;7(5):591–605.

17. Nobles J, Fox C, Inman-Ward A, Beasley T, Redwood S, Jago R, et al. Navigating the river(s) of systems change: a multi-methods, qualitative evaluation exploring the implementation of a systems approach to physical activity in Gloucestershire, England. BMJ Open. 2022;12(8).

18. Rosas S, Knight E. Evaluating a complex health promotion intervention: case application of three systems methods. Critical Public Health. 2019;29(3):337–52.

19. Hall J, Bingham DD, Seims A, Dogra SA, Burkhardt J, Nobles J, et al. A whole system approach to increasing children’s physical activity in a multi-ethnic UK city: a process evaluation protocol. BMC Public Health. 2021;21(1).

20. Hanson S, Jones A. Missed opportunities in the evaluation of public health interventions: a case study of physical activity programmes. BMC Public Health. 2017;17(1).

21. Egan M, McGill E, Penney TL, Cuevas RAd, Er V, Orton L, et al., editors. NIHR SPHR Guidance on Systems Approaches to Local Public Health Evaluation. Part 1: Introducing systems thinking 2019.

22. Nau T, Bauman A, Smith BJ, Bellew W. A scoping review of systems approaches for increasing physical activity in populations. Health Research Policy and Systems. 2022;20(1).

23. Mcgill E, Marks D, Er V, Penney T, Petticrew M, Egan M. Qualitative process evaluation from a complex systems perspective: A systematic review and framework for public health evaluators. PLOS Medicine. 2020;17(11).

24. Breslin G, Wills W, Bartington S, Bontoft C, Fakoya O, Freethy I, et al. Evaluation of a whole system approach to diet and healthy weight in the east of Scotland: Study protocol. PLOS ONE. 2022;17(3).

25. UN General Assembly, editor Convention on the Rights of the Child 20 November 1989.

26. Hoekstra F, Mrklas KJ, Khan M, Mckay RC, Vis-Dunbar M, Sibley KM, et al. A review of reviews on principles, strategies, outcomes and impacts of research partnerships approaches: a first step in synthesising the research partnership literature. Health Research Policy and Systems. 2020;18(1).

27. Vohland K, Land-Zandstra A, Ceccaroni L, Lemmens R, Perelló J, Ponti M, et al. The Science of Citizen Science Evolves. Chapter 1 in Vohland, K et al (Eds)(2021) The Science of Citizen Science Springer https://doiorg/101007/978-3-030-58278-4 xpp 1-12. 2021.

28. Rüfenacht S, Woods T, Agnello G, Gold M, Hummer P, Land-Zandstra A, et al. Communication and Dissemination in Citizen Science. 2021. p. 475–94.

29. Warren CA, Marciano JE. Activating student voice through Youth Participatory Action Research (YPAR): policy-making that strengthens urban education reform. International Journal of Qualitative Studies in Education (QSE). 2018;31(8):684–707.

30. Martinez LS, Yan CT, McClay C, Varga S, Zaff JF. Adult reflection on engaging youth of color in research and action: A case study from five US Cities. Journal of Adolescent Research. 2020;35(6):699–727.

31. Lane HG, Porter KJ, Hecht E, Harris P, Zoellner JM. A Participatory Process to Engage Appalachian Youth in Reducing Sugar-Sweetened Beverage Consumption. Health Promotion Practice. 2019;20(2):258–68.

32. Bender K, Barman-Adhikari A, DeChants J, Haffejee B, Anyon Y, Begun S, et al. Asking for Change: Feasibility, acceptability, and preliminary outcomes of a manualized photovoice intervention with youth experiencing homelessness. Children and Youth Services Review. 2017;81:379–89.

33. Ozer EJ, Douglas L. The Impact of Participatory Research on Urban Teens: An Experimental Evaluation. American Journal of Community Psychology. 2013;51(1-2):66–75.

34. Ballard HL, Dixon CGH, Harris EM. Youth-focused citizen science: Examining the role of environmental science learning and agency for conservation. Biological Conservation. 2017;208:65–75.

35. Cuevas-Parra P. Co-Researching With Children in the Time of COVID-19: Shifting the Narrative on Methodologies to Generate Knowledge. International Journal of Qualitative Methods. 2020;19:160940692098213.

36. Bälter K, Rydenstam T, Fell T, King AC, Buli BG. Data from an Our Voice citizen science initiative in neighborhoods with low socioeconomic status in Sweden: A proof of concept for collecting complex data. Data in brief. 2020;33.

37. ECSA (European Citizen Science Association). 10 Principles of Citizen Science. https://ecsa.citizen-science.net/wp-content/uploads/2021/05/ECSA_Ten_Principles_of_CS_English.pdf: ECSA (European Citizen Science Association); 2015. Accessed 10.10.22

38. Maker Castro E, López Hernández G, Karras-Jean Gilles J, Novoa A, The New Generation C, Suárez-Orozco C. “Everyone collaborated and came together”: The civic promise (and pitfalls) of yPAR for immigrant-origin students in an era of deportation. Cultur Divers Ethnic Minor Psychol. 021.

39. Scott MA, Pyne KB, Means DR. Approaching praxis: YPAR as critical pedagogical process in a college access program. The High School Journal. 2015;98(2):138–57.

40. Wagaman MA, Sanchez I. Looking through the magnifying glass: A duoethnographic approach to understanding the value and process of participatory action research with LGBTQ youth. Qualitative Social Work. 2016;16(1):78–95.

41. Kieslinger B, Schürz, Stefanie, Mayer, Katja, & Schäfer, Teresa. CoActD7.2: Interim Impact Assessment Report.. Zenodo.; 2021.

42. Kieslinger B, Schaefer T, Heigl F, Dörler D, Richter A, Bonn A. The Challenge of Evaluation: An Open Framework for Evaluating Citizen Science Activities. 2017.

43. Bradford Council. Demographics of Bradford District. In: Assessment JSN, District TPoB, editors. 2020. Accessed 10.10.22

44. Office for National Statistics. What are the regional differences in income and productivity? In: Statistics OfN, editor. 2021.

45. (ISPAH). International Society for Physical Activity and Health. ISPAH’s Eight Investments That Work for Physical Activity. 2020.

46. Dogra SA, Rai KK, Barber SE, McEachan RRC, Adab P, Sheard L. Delivering a childhood obesity prevention intervention using Islamic religious settings in the UK: What is most important to the stakeholders? Preventive Medicine Reports. 2021;22.

47. Daly-Smith A, Quarmby T, Archbold VSJ, Corrigan N, Wilson D, Resaland GK, et al. Using a multi-stakeholder experience-based design process to co-develop the Creating Active Schools Framework. International Journal of Behavioral Nutrition and Physical Activity. 2020;17(1).

48. Westhorp G. Using complexity-consistent theory for evaluating complex systems. Evaluation. 2012;18(4):405–20.

49. Bird PK, Mceachan RRC, Mon-Williams M, Small N, West J, Whincup P, et al. Growing up in Bradford: protocol for the age 7–11 follow up of the Born in Bradford birth cohort. BMC Public Health. 2019;19(1).

50. Bingham D. The effectiveness and health impact of a whole-systems physical activity intervention at increasing the physical activity levels of children aged 5-11 years. In Preperation

51. Spasiano A, Grimaldi S, Braccini AM, Nardi F. Towards a Transdisciplinary Theoretical Framework of Citizen Science: Results from a Meta-Review Analysis. Sustainability. 2021;13(14):7904.

52. Kullenberg C, Kasperowski D. What Is Citizen Science? – A Scientometric Meta-Analysis. PLOS ONE. 2016;11(1):e0147152.

53. Haklay M, Dörler D, Heigl F, Manzoni M, Hecker S, Vohland K. What Is Citizen Science? The Challenges of Definition. The Science of Citizen Science: Springer International Publishing; 2021.. 13–33.

54. Cooper CBL, Bruce V. The Rightful Place of Science: Citizen Science 2016.

55. Haklay M. Citizen Science and Volunteered Geographic Information: Overview and Typology of Participation. 2013. p. 105–22.

56. Schäfer T, Kieslinger B. Supporting emerging forms of citizen science: a plea for diversity, creativity and social innovation. Journal of Science Communication. 2016;15(02):Y02.

57. Shirk JL, Ballard HL, Wilderman CC, Phillips T, Wiggins A, Jordan R, et al. Public Participation in Scientific Research: a Framework for Deliberate Design. Ecology and Society. 2012;17(2).

58. Cohen DK, Heha Bhandari; Stewert, David; Rees, Nicholas. Advocacy toolkit: A guide to influencing decisions that improve childrens lives. https://www.susana.org/en/knowledge-hub/resources-and-publications/library/details/2442: United Nations Childres Fund (UNICEF); 2010. Accessed 10.10.22

59. Islam S AA, Haklay M & McEachan R. Co-production in ActEarly: nothing about us without us.: Bradford Institute for Health Research & University College London; 2022.

60. Lewis J. Analysing Qualitative Longitudinal Research in Evaluations. Social Policy and Society. 2007;6(4):545–56.

61. Neale B. Qualitative Longitudinal Research, Research Methods. London: Bloomsbury Publishing; 2019.

62. Atkin AJ, Sharp SJ, Harrison F, Brage S, Van Sluijs EM. Seasonal Variation in Children’s Physical Activity and Sedentary Time. Med Sci Sports Exerc. 2016;48(3):449–56.

63. Shaw C, Brady L-M, Davey C. NCB Guidelines for Research With Children and Young People 2011.

64. Adler K, Salanterä S, Zumstein-Shaha M. Focus Group Interviews in Child, Youth, and Parent Research: An Integrative Literature Review. International Journal of Qualitative Methods. 2019;18:160940691988727.

65. Terras MM, Ramsay J. Family Digital Literacy Practices and Children’s Mobile Phone Use. Front Psychol. 2016;7:1957-.

66. (NSPCC) NSftPoCtC. Keeping children safe away from home. 2022 [Available from: https://www.nspcc.org.uk/keeping-children-safe/away-from-home/at-school/. Accessed 10.1022

67. Ofcom. Children and Parents: Media Use and Attitudes Report 2018. 2019.

68. Brown HE, Atkin AJ, Panter J, Wong G, Chinapaw MJM, Van Sluijs EMF. Family-based interventions to increase physical activity in children: a systematic review, meta-analysis and realist synthesis. Obesity Reviews. 2016;17(4):345–60.

69. Ungar WJ, Mirabelli C, Cousins M, Boydell KM. A qualitative analysis of a dyad approach to health-related quality of life measurement in children with asthma. Social Science & Medicine. 2006;63(9):2354–66.

70. Yardley L, Ainsworth B, Arden-Close E, Muller I. The person-based approach to enhancing the acceptability and feasibility of interventions. Pilot and Feasibility Studies. 2015;1(1).

71. Bellg A, Borrelli B, Resnick B, Hecht J, Minicucci D, Ory M, et al. Enhancing Treatment Fidelity in Health Behavior Change Studies: Best Practices and Recommendations From the NIH Behavior Change Consortium. Health psychology : official journal of the Division of Health Psychology, American Psychological Association. 2004;23:443–51.

72. Lawton R, Mceachan R, Jackson C, West R, Conner M. Intervention fidelity and effectiveness of a UK worksite physical activity intervention funded by the Bupa Foundation, UK. Health Promot Int. 2015;30(1):38–49.

73. Allen J, Linnan L, Emmons K. Fidelity and Its Relationship to Implementation Effectiveness, Adaptation, and Dissemination. 2012. p. 281–304.

74. Sekhon M, Cartwright M, Francis JJ. Acceptability of healthcare interventions: an overview of reviews and development of a theoretical framework. BMC Health Services Research. 2017;17(1).

75. Moore GF, Audrey S, Barker M, Bond L, Bonell C, Hardeman W, et al. Process evaluation of complex interventions: Medical Research Council guidance. BMJ. 2015;350(mar19 6):h1258–h.

76. Noonan RJ, Boddy LM, Fairclough SJ, Knowles ZR. Write, draw, show, and tell: a child-centred dual methodology to explore perceptions of out-of-school physical activity. BMC Public Health. 2016;16(1).

77. Ritchie JS L. Qualitative data analysis for applied pollicy research. IN Analyzing qualitative data In: R.G. Bab, editor. Abingdon: Routledge; 1994. p. 173–94.

78. Akom A, Shah A, Nakai A, Cruz T. Youth participatory action research (YPAR) 2.0: How technological innovation and digital organizing sparked a food revolution in East Oakland. International Journal of Qualitative Studies in Education. 2016;29(10):1287–307.

79. (RCPCH) Recipes for Engagement: Children and young people in the lead. In: (RCPCH). https://wwwrcpchacuk/sites/default/files/2018-09/recipes_for_engagement_2018pdf. The Royal College of Paediatrics and Child Health (RCPCH); 2018. Accessed 10.10.22

80. Genuis SK, Willows N, Jardine CG. Partnering with Indigenous student co-researchers: improving research processes and outcomes. International journal of circumpolar health. 2015;74:27838.

81. Grossoehme D, Lipstein E. Analyzing longitudinal qualitative data: the application of trajectory and recurrent cross-sectional approaches. BMC Research Notes. 2016;9(1).

82. Braun V, Clarke V. Using thematic analysis in psychology. Qualitative Research in Psychology. 2006;3(2):77–101.

83. Little S. Rivers of multilingual reading: exploring biliteracy experiences among 8-13-year old heritage language readers. Journal of Multilingual and Multicultural Development. 2021:1–14.

84. Thomson R, Holland J. Hindsight, foresight and insight: The challenges of longitudinal qualitative research. International Journal of Social Research Methodology. 2003;6(3):233–44.

85. Schaefer T, Kieslinger B, Brandt M, Bogaert V. Evaluation in Citizen Science: The Art of Tracing a Moving Target. 2021. p. 495–514.

86. Nixon LS, Hudson N, Culley L, Lakhanpaul M, Robertson N, Johnson MRD, et al. Key considerations when involving children in health intervention design: reflections on working in partnership with South Asian children in the UK on a tailored Management and Intervention for Asthma (MIA) study. Res Involv Engagem. 2022;8(1):9.

87. Nabors LA, Ramos V, Weist MD. Use of Focus Groups as a Tool for Evaluating Programs for Children and Families. Journal of educational and psychological consultation. 2001;12(3):243–56.

88. Canosa A, Graham A, Wilson E. Reflexivity and ethical mindfulness in participatory research with children: What does it really look like? Childhood: A Global Journal of Child Research. 2018;25(3):400–15.

89. Wartenweiler D, Mansukhani R. Participatory Action Research with Filipino Street Youth: Their Voice and Action against Corporal Punishment. Child Abuse Review. 2016;25(6):410–23.

90. Halliday AJ, Kern ML, Garrett DK, Turnbull DA. The student voice in well-being: a case study of participatory action research in positive education. Educational Action Research. 2019;27(2):173–96.

91. Rasmussen LM, Cooper C. Citizen Science Ethics. Citizen Science: Theory and Practice. 2019;4(1).

92. Haklay MM, A.; Balázs, B.; Kieslinger, B.; Greshake Tzovaras, B.; Nold, C.; Dörler, D.; Fraisl, D.; Riemenschneider, D.; Heigl, F,. The ECSA Characteristics of Citizen Science: ECSA; 2020.

93. Paleco C, García Peter S, Salas Seoane N, Kaufmann J, Argyri P. Inclusiveness and Diversity in Citizen Science. The Science of Citizen Science: Springer International Publishing; 2021. p. 261–81.

